# Accounting for uncertainty in the expected treatment effect substantially increases the sample size required for randomised trials: implications for the feasibility of clinical trials in anaesthesia and critical care

**DOI:** 10.64898/2026.06.19.26356097

**Authors:** David Sidebotham, C. Jake Barlow

## Abstract

**Background:** Multicentre trials in anaesthesia and critical care report low rates of statistically significant differences. This finding may partly reflect conventional sample size methods, which assume a fixed treatment effect. Assurance methods use a design prior to represent uncertainty in the expected treatment effect, which may provide a more realistic way of estimating sample sizes.

**Methods:** We calculated power curves across a range of effect sizes, design priors, and sample sizes using frequentist and Bayesian assurance methods and compared the sample sizes required to achieve 80% and 90% power to the conventional method. We standardised the design priors across effect sizes using the coefficient of variation (CV). We derived a theoretical limit for achievable power. We validated a normal approximation to the Bayesian posterior distribution.

**Results:** Frequentist and Bayesian assurance methods produced similar power curves across all scenarios. At a CV of 0.5 – reflecting realistic prior uncertainty in the expected effect size – both methods required sample sizes that were approximately 1.5 to 3.5 times larger than the conventional method. The theoretical power limit depends only on the CV of the design prior and holds true across all effect sizes. The normal approximation to the Bayesian posterior distribution matched the results obtained from Markov chain Monte Carlo sampling.

**Conclusions:** Incorporating clinical uncertainty in the expected effect size substantially increases the sample size required to achieve adequate power, which has important implications for the feasibility of randomised trials in anaesthesia and critical care.

## Introduction

A common design for randomised trials in anaesthesia and critical care is a two-arm study where the outcome is a binary event. Despite Bayesian inference offering several advantages,^1^ null hypothesis significance testing (NHST) remains the dominant statistical methodology.

When estimating the sample size for a randomised trial, researchers choose the expected event rates in the control and intervention groups, the design power (typically 80-90%), and the significance threshold (alpha [α], typically 0.05).^2^ There is compelling evidence that researchers routinely overestimate the plausible effect size.^3–7^ Overestimating the expected effect size means the achieved power of the study will be less than the design power, increasing the chance of a type II error (failing to reject the null hypothesis when it is false). Low power also means that on the rare occasions when a result is statistically significant the observed effect size is an overestimate of the true effect size, a phenomenon known as the winner’s curse.^8^

Even researchers who are aware of the danger of inflating the expected effect size face a fundamental challenge: genuine uncertainty about the effect size. Indeed, this uncertainty is the very reason the trial is being done. Conventional methods take no account of uncertainty and estimate the sample size assuming the effect size is fixed. Conventional sample size calculations are often called conditional power methods because the calculation is conditional on the assumed effect size being the true effect size. By contrast, assurance methods quantify uncertainty in the expected effect size through a design prior and calculate the sample size by averaging (integrating) power across the range of plausible effect sizes.^9^

Assurance methods can be used with both frequentist (i.e., NHST) and Bayesian inference. In both cases, researchers specify a success criterion. The proportion of the time the criterion is satisfied at a particular sample size becomes the assurance power at that sample size. For frequentist assurance, the success criterion for a single trial is of the form *P* ≤ *α*. For Bayesian assurance, the success criterion for a single trial is typically that the posterior probability the treatment is beneficial exceeds a threshold; for example, 97.5%. Thus, a Bayesian success criterion might be of the form *P*(*OR* < 1|data) > 0.975, where OR is the odds ratio and the vertical operator means ‘given that’.

A recent correspondence in the journal by Markus Huber demonstrated that incorporating uncertainty about the expected treatment effect through a frequentist assurance method substantially reduces the expected probability of trial success compared to conventional power calculations.^10^ In this article, we extend Huber’s work to both frequentist and Bayesian assurance methods, and characterise the sample sizes required to achieve 80% and 90% assurance power across a range of effect sizes and design priors. We also derive a theoretical power limit that depends only on the coefficient of variation (CV) of the design prior. To make the Bayesian assurance method computationally tractable, we use (and validate) a normal approximation to the Bayesian posterior distribution that reproduces the results obtained from Markov chain Monte Carlo (MCMC) sampling at a fraction of the computational cost.

## Methods

A detailed description of the methods is provided in Supplementary Material 1 (SM1). All simulations were done in R.^11^ The R code used for the simulations is available at https://github.com/cjdbarlow/papers.

### Scenarios

We considered a two-arm randomised trial with a binary outcome where the expected control group event rate was 50%. We specified four effect sizes by setting the expected intervention event rate to 40%, 45%, 48%, and 49%, giving expected risk differences (RDs) of –10%, –5.0%, –2.0%, and –1.0%, respectively (expected ORs of 0.67, 0.82, 0.92, 0.96, respectively). For each effect size, we calculated power curves using three methods: (1) a conventional frequentist power formula, (2) a frequentist assurance method, and (3) a Bayesian assurance method, referred to henceforth as ‘Bayesian power’.

### Statistical model

We used logistic regression as the statistical model, defined as:

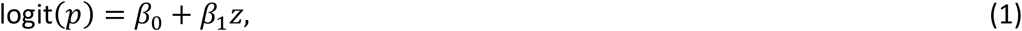

where *p* is the event probability, logit(*p*) is the logit function for *p* (defined as log(*p*⁄(1 − *p*)), *β*_0_ is the intercept coefficient, *β*_1_ is the slope coefficient, and *z* is an indicator variable that takes the value 1 when a patient is a member of the intervention group and 0 when a patient is a member of the control group. The coefficient *β*_0_ represents the log odds the event in the control group and the coefficient *β*_1_ represents the log OR of the event in the intervention group relative to the control group.

### Prior uncertainty: the design priors

For each expected effect size, we specified 10 normal (Gaussian) design priors on β_1,_ representing different degrees of certainty in the treatment effect. To ensure the prior standard deviation (SD) was comparable across the four effect sizes, we specified a constant coefficient of variation (CV) for each design prior, defined as:

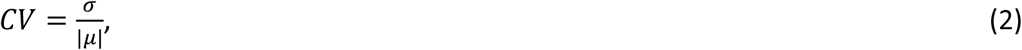

where |μ| is the absolute value of the prior mean and σ is the prior SD. We specified CVs of 0.1, 0.2, 0.3, 0.4, 0.5, 0.75, 1.0, 1.19, 1.5, and 2.0, reflecting progressively reduced certainty in the true treatment effect. Specifying a constant CV across effect sizes is equivalent to holding a constant prior belief that the intervention is beneficial. The CV of 1.19 was specifically included, as it represents the theoretical threshold below which 80% power remains achievable (see below). Specifying a constant CV imposes different SDs across the priors for the four effect sizes. For instance, for a RD of –10% (OR 0.67, log OR –0.40), a CV of 0.5 imposes an SD of 0.2 on the log scale whereas for a RD of –1.0% (OR 0.96, log OR –0.04) a CV of 0.5 imposes a SD of 0.02 on the log scale. Tables SM11.1 and SM11.2 summarise the design priors across effect sizes and CVs.

For all simulations, we specified a normal prior on β_0_ with mean 0 and SD 0.1. The prior on β_0_ centres the normal distribution tightly on the expected control event rate of 50% (log odds 0).

### Conventional (conditional) power

For each effect size, CV, and sample size, we calculated conventional power using Equation SM1.6, which is the standard power formula for two equal groups and a binary outcome at a one-sided significance test for α=0.025 where the expected control event rate is 50%.

### Frequentist assurance power (see SM1.4)

We calculated frequentist assurance using random draws from the β_1_ design prior and inputted the values into the Equation SM1.9. The success criterion was a one-sided P-value ≤0.025. For each effect size, design prior, and sample size we drew 5,000 random samples and averaged power across the samples (Monte Carlo integration). Samples where β_1_≥0 (representing neutral or harmful effects) were assigned 0 power, ensuring directional consistency with the Bayesian success criterion (see below). Table SM1.3 shows the sample sizes and steps for the frequentist assurance calculations.

### Bayesian assurance power (see SM1.5)

We calculated Bayesian power by drawing random sample pairs from β_0_ and β_1_ for the design priors. We converted the regression coefficients into event probabilities and simulated data for a randomised trial. For each simulated trial at each effect size, design prior, and sample size, we fitted a Bayesian logistic regression model and estimated the posterior value for β_1_ (i.e., the posterior log OR). To fit the models, we specified identical normal analysis priors^*^ on β0 and β1 with mean 0 and SD 0.5. The analysis prior on β0 is centred on a control group event rate of 50% (log odds of 0) and is weakly informative. The analysis prior on β_1_ is neutral (centred on a log OR of 0) and is weakly informative. We specified a Bayesian success criterion of *P*(*β*_1_ < 0|data) > 0.975, which is equivalent to *P*(log(*OR*) < 0|data) > 0.975 and *P*(*OR* < 1|data) > 0.975. Bayesian power was defined as the proportion of simulated trials at each sample size that satisfied the success criterion.

We used a normal approximation method to compute the posterior distribution for β_1_, as described in SM1.8. We validated the normal approximation using full MCMC sampling on a representative subset of simulations (see SM1.9). Table SM1.4 summarises the sample sizes and steps for the Bayesian power calculations.

### Theoretical power limit

We derived the theoretical ceiling for frequentist assurance and Bayesian power.

## Results

Figures 1-4 show the power curves for frequentist and Bayesian assurance power for the design priors across the four effect sizes. Table 1 shows the theoretical power limit and the sample sizes required to achieve 80% and 90% power for each design prior. The assurance power at different sample sizes is shown in Supplementary Material 2.

**Figure 1.**
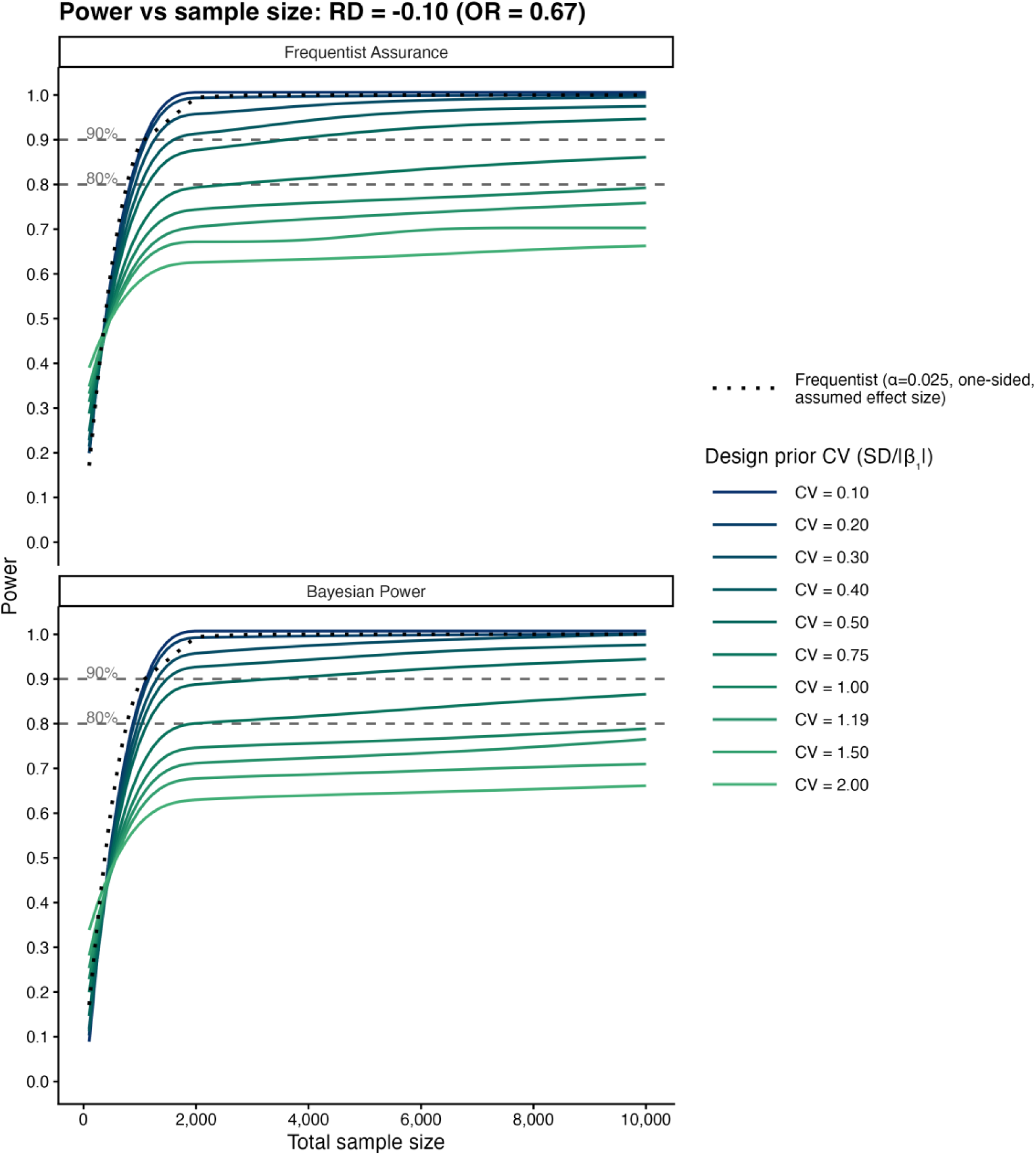
Power curves for an expected risk difference (RD) of –10% at different coefficients of variation (CV) applied to the design prior on β_1_. The top panel shows the results for frequentist assurance and the bottom panel shows the results for Bayesian assurance power. The power curve for a conventional frequentist power calculation (one-sided alpha of 0.025) is shown as a black dashed line.

**Figure 2.**
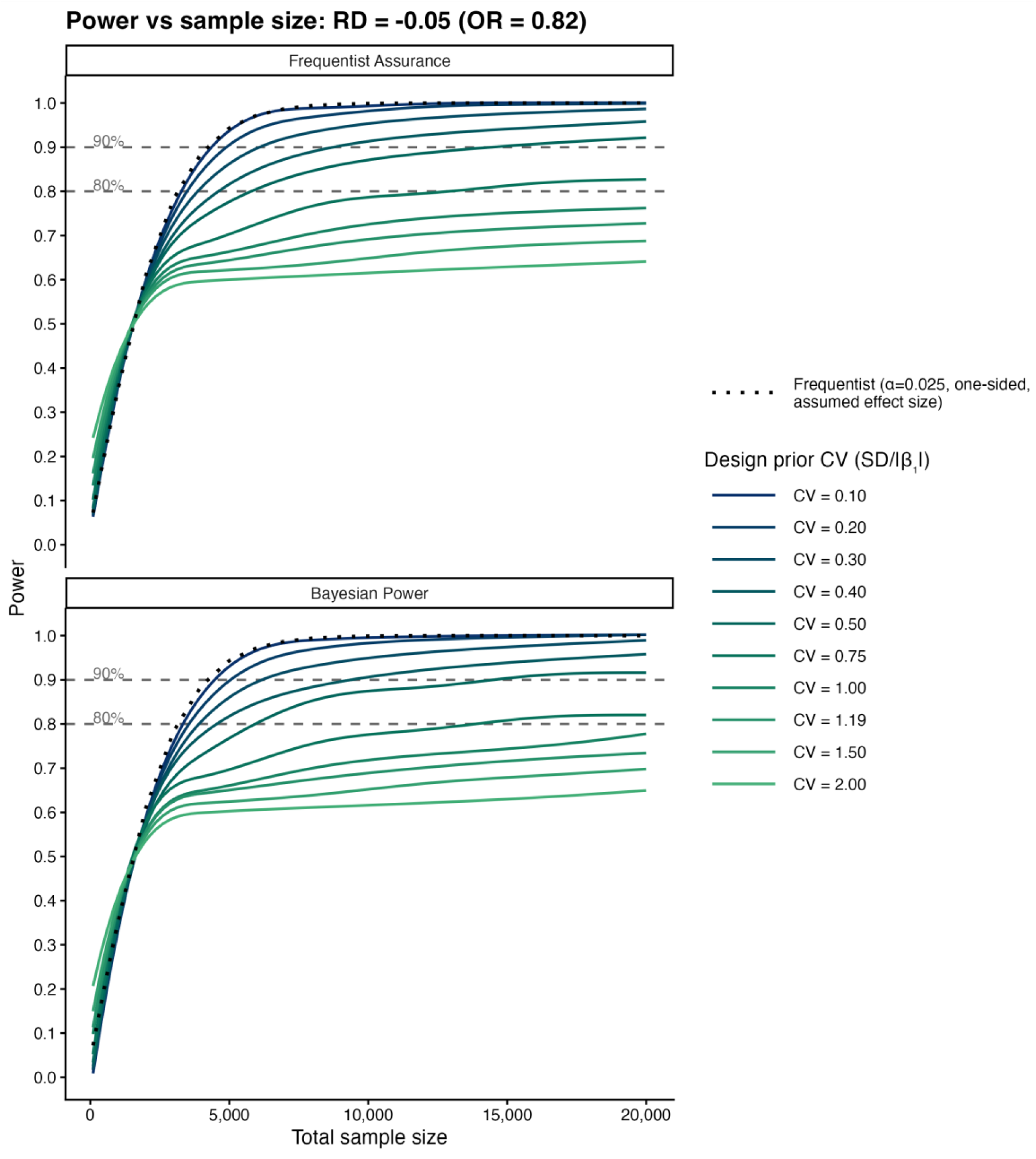
Power curves for an expected risk difference (RD) of –5.0% at different coefficients of variation (CV) applied to the design prior on β_1_. The top panel shows the results for frequentist assurance and the bottom panel shows the results for Bayesian assurance power. The power curve for a conventional frequentist power calculation (one-sided alpha of 0.025) is shown as a black dashed line.

**Figure 3.**
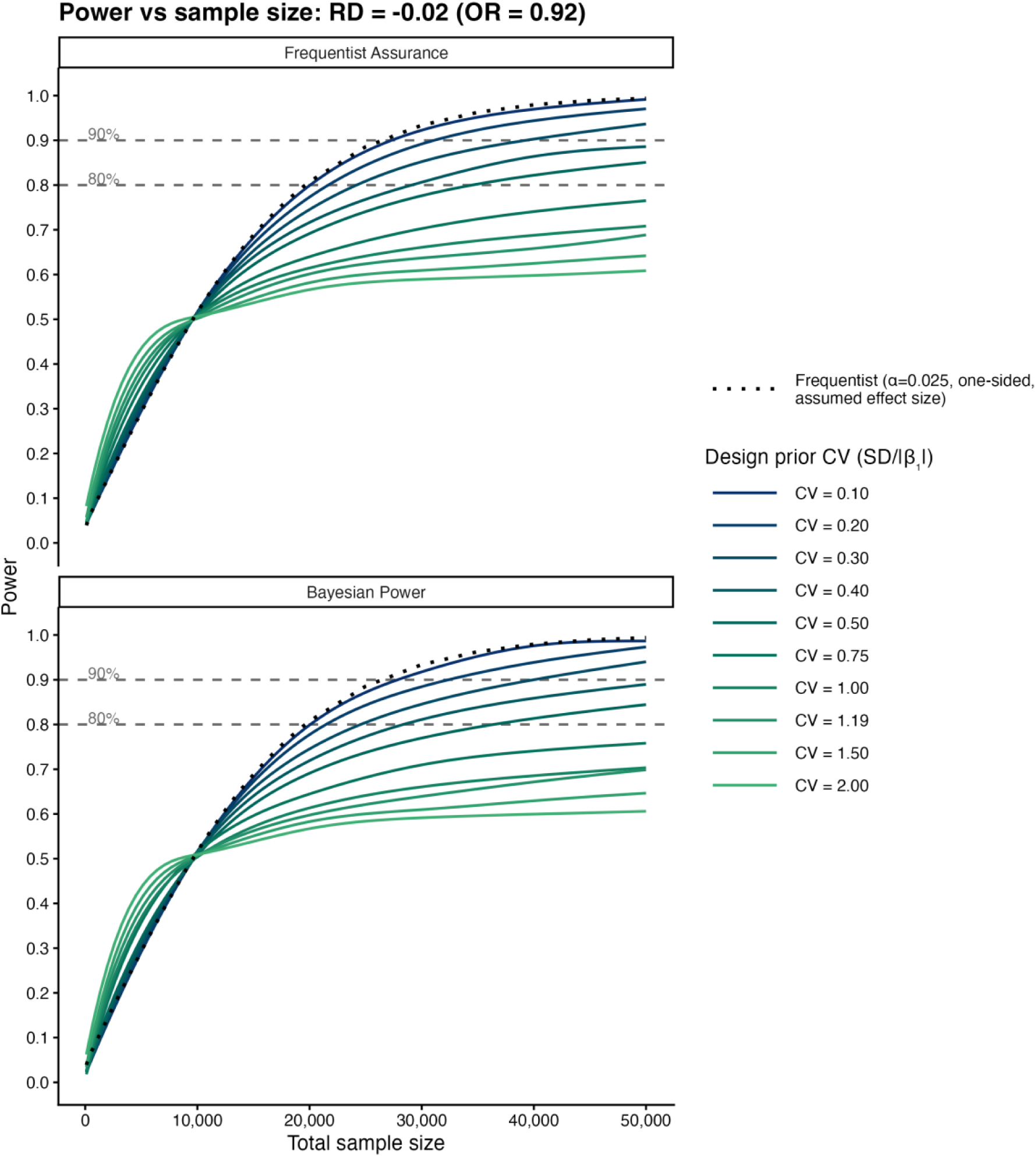
Power curves for an expected risk difference (RD) of –2.0% at different coefficients of variation (CV) applied to the design prior on β_1_. The top panel shows the results for frequentist assurance and the bottom panel shows the results for Bayesian assurance power. The power curve for a conventional frequentist power calculation (one-sided alpha of 0.025) is shown as a black dashed line.

**Figure 4.**
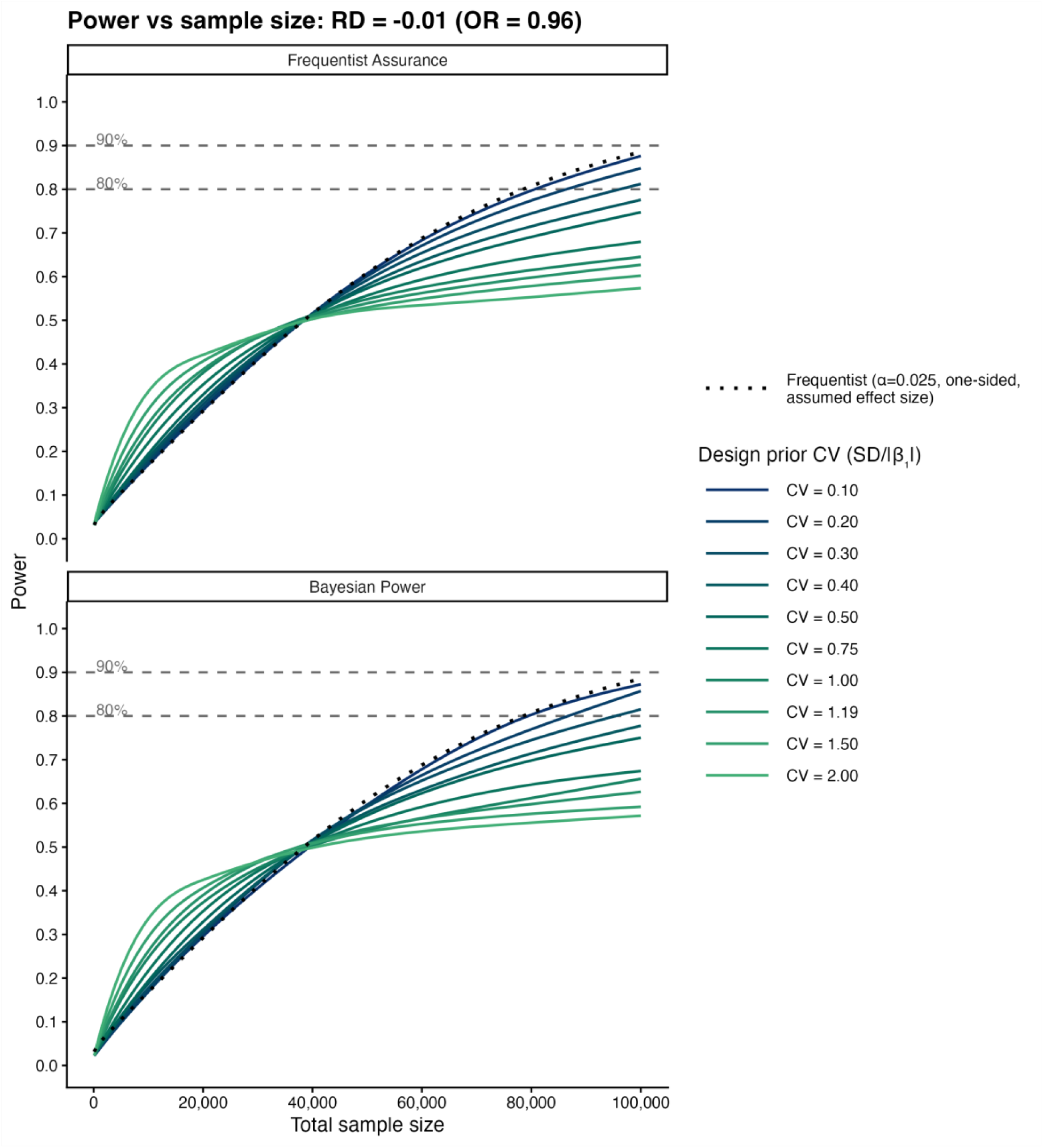
Power curves for an expected risk difference (RD) of –1.0% at different coefficients of variation (CV) applied to the design prior on β_1_. The top panel shows the results for frequentist assurance and the bottom panel shows the results for Bayesian assurance power. The power curve for a conventional frequentist power calculation (one-sided alpha of 0.025) is shown as a black dashed line.

**Table 1.** Sample size to achieve 80% and 90% assurance power by effect size and method (frequentist, Bayesian) for the design priors on β_1_. Empty cells indicate that the sample sized exceeded the upper limit of the simulated sample size. CV indicates the coefficient of variation of the design prior on β_1_. power_∞_ denotes the maximum theoretical power as *n* → ∞. OR indicates odds ratio and log OR is the natural logarithm of the odds ratio.

|  |  |  |  |  |  |
| --- | --- | --- | --- | --- | --- |
| **Expected risk difference -10% (OR 0.67, log OR -0.41)**<br>Conventional frequentist sample size: 774 (80% power), 1037(90% power)<br>Maximum sample size: 20,000. |  |  |  |  |  |
|  | **Frequentist assurance** |  | **Bayesian assurance** |  |  |
| **CV** | **80% power** | **90% power** | **80% power** | **90% power** | **power<sub>∞</sub>** |
| 0.10 | 787 | 1,179 | 848 | 1,233 | 1.00 |
| 0.20 | 845 | 1,399 | 912 | 1,457 | 1.00 |
| 0.30 | 946 | 1,705 | 1,000 | 1,803 | 1.00 |
| 0.40 | 1,161 | 2,235 | 1,296 | 1,976 | 0.99 |
| 0.50 | 1,530 | 3,509 | 1,439 | 3,595 | 0.98 |
| 0.75 | 3,188 | - | 3,216 | - | 0.91 |
| 1.00 | 15,426 | - | 17,040 | - | 0.84 |
| 1.19 | - | - | - | - | 0.80 |
| 1.50 | - | - | - | - | 0.75 |
| 2.00 | - | - | - | - | 0.69 |
| **Expected risk difference -5% (OR 0.82, log OR -0.20)**<br>Conventional frequentist sample size: 3,126 (80% power), 4,232 (90% power).<br>Maximum sample size 50,000. |  |  |  |  |  |
|  | **Frequentist assurance** |  | **Bayesian assurance** |  |  |
| **CV** | **80% power** | **90% power** | **80% power** | **90% power** | **power<sub>∞</sub>** |
| 0.10 | 3,238 | 4,416 | 3,361 | 4,491 | 1.00 |
| 0.20 | 3,493 | 4,965 | 3,611 | 4,950 | 1.00 |
| 0.30 | 3,820 | 6,218 | 3,838 | 6,250 | 1.00 |
| 0.40 | 4,585 | 8,758 | 4,531 | 9,667 | 0.99 |
| 0.50 | 5,729 | 14,113 | 5,391 | 14,778 | 0.98 |
| 0.75 | 12,926 | - | 12,841 | - | 0.91 |
| 1.00 | - | - | 49,333 | - | 0.84 |
| 1.19 | - | - | - | - | 0.80 |
| 1.50 | - | - | - | - | 0.75 |
| 2.00 | - | - | - | - | 0.69 |
| **Expected risk difference -2.0% (OR 0.92, log OR -0.08)**<br>Conventional frequentist sample size: 19,641 (80% power), 26,324 (90% power).<br>Maximum sample size: 300,000. |  |  |  |  |  |
|  | **Frequentist assurance** |  | **Bayesian assurance** |  |  |

| CV | 80% power | 90% power | 80% power | 90% power | power<sub>∞</sub> |
| --- | --- | --- | --- | --- | --- |
| 0.10 | 20,085 | 27,668 | 20,736 | 28,177 | 1.00 |
| 0.20 | 21,913 | 31,346 | 21,531 | 32,470 | 1.00 |
| 0.30 | 24,283 | 39,635 | 24,338 | 39,141 | 1.00 |
| 0.40 | 29,222 | 55,860 | 28,544 | 54,271 | 0.99 |
| 0.50 | 34,534 | 93,724 | 36,746 | 90,952 | 0.98 |
| 0.75 | 77,926 | - | 93,182 | - | 0.91 |
| 1.00 | 278,939 | - | 286,667 | - | 0.84 |
| 1.19 | - | - | - | - | 0.80 |
| 1.50 | - | - | - | - | 0.75 |
| 2.00 | - | - | - | - | 0.69 |
| **Expected risk difference -1.0% (OR 0.96, log OR -0.04)**<br>Conventional frequentist sample size: 78,529 (80% power).<br>Maximum sample size: 300,000. |  |  |  |  |  |
|  | Frequentist assurance |  | Bayesian assurance |  |  |
| CV | 80% power | 90% power | 80% power | 90% power | power<sub>∞</sub> |
| 0.10 | 80,405 | 109,717 | 80,100 | 108,082 | 1.00 |
| 0.20 | 86,157 | 124,240 | 86,761 | 127,674 | 1.00 |
| 0.30 | 96,290 | 155,481 | 96,556 | 155,510 | 1.00 |
| 0.40 | 113,820 | 216,514 | 115,122 | 239,000 | 0.99 |
| 0.50 | 138,591 | - | 137,818 | - | 0.98 |
| 0.75 | 295,025 | - | 294,773 | - | 0.91 |
| 1.00 | - | - | - | - | 0.84 |
| 1.19 | - | - | - | - | 0.80 |
| 1.50 | - | - | - | - | 0.75 |
| 2.00 | - | - | - | - | 0.69 |

The theoretical power ceiling (power_∞_) as *n* → ∞ was derived as:

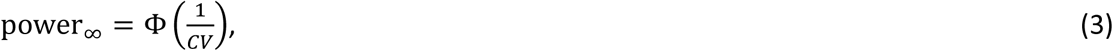

where Φ(.) indicates the cumulative distribution function of the standard normal distribution (see SM1.10). The theoretical power limit equates to the prior probability the intervention is beneficial (Table SM1.2). The CV thresholds for the design priors at which 80% and 90% power are still theoretically achievable are 1.19 and 0.74, respectively.

The normal approximation to the Bayesian posterior distribution was robust across a range of effect sizes and design priors (SM1.9, Fig SM1.1).

## Discussion

Our findings show that incorporating uncertainty in the expected effect size through a design prior increases the sample size that is required to achieve power of 80% or 90% compared to the conventional method. Only when prior confidence in the effect size is extremely high (CV 0.1-0.3, Tables SM1.1 and SM1.2) are the assurance power curves similar to the conventional method. When the CV of the design prior is 0.5, the sample size required to achieve assurance power of 80% or 90% is roughly 1.5 to 3.5 times greater than the sample size obtained from the conventional method. A CV of 0.5 represents a strong belief that the treatment is beneficial but accommodates genuine uncertainty regarding the effect size (Table SM1.2).

Consider a hypothetical multicentre trial targeting an intervention for sepsis where the expected control group mortality is 50%. The researchers think the most likely mortality-sparing effect of the intervention is a RD of –10% but consider it might plausibly vary between –19% and +2%, corresponding to a design prior with a CV of 0.5.^†^ Using the conventional method, the sample size required to achieve 80% power is 771 (Table 1). However, if the researchers accommodate their uncertainty as a design prior, then, with the frequentist assurance method, the sample size increases to 1,530. Now, consider an intervention for sepsis where the researchers think the most likely mortality-sparing effect is a RD of –5% with a plausible range between –9.8% and +1%, which also corresponds to a design prior with a CV of 0.5. Using the conventional method, the sample size required to achieve 80% power is 3,156 but to achieve 80% frequentist assurance power requires a sample size of 5,729. Failure to account for this uncertainty in the effect size potentially means achieved power of a trial is less than the design power, which may partly explain the low proportion of multicentre trials in anaesthesia and critical care that report statistically significant differences.^3, 4, 7^

The usual frequentist success criterion is a two-sided P-value at α=0.05. When specifying a design prior where most of the probability mass is in the region of benefit – as in our simulations, where the probability of benefit ranged from almost 100% (CV 0.1) to 69% (CV 2.0) –a one-sided success criterion at the 0.025 threshold is numerically equivalent to a two-sided test at the 0.05 threshold (see SM1.6).

We derived a theoretical ceiling on achievable power (Equation 3) that depends only on the CV of the design prior and remains constant across effect sizes and inferential methodologies (see SM1.10). Our finding is concordant with the theoretical work of De Santis, Gubbiotti, and Brutti, who identified a ceiling on power that depends only on the design prior.^12, 13^ In SM1.11 we show that Equation 3 is mathematically identical to the result obtained by De Santis and Gubbiotti.^12^ We also demonstrated that the theoretical limit on power is equivalent to the prior probability the treatment is beneficial (see SM1.10, Table SM1.1).

Our findings for frequentist assurance power are in broad agreement with Huber’s method despite substantial differences between the two methodologies.^10^ Huber specified a uniform design prior bounded between zero effect and the assumed effect size, which has the advantage of simplicity but may not fully capture the range of plausible treatment effects. Because Huber’s uniform prior places zero probability mass on harmful effects, the theoretical ceiling on power is always 100%. By contrast, we specified normal distributions that place at least some probability mass on harmful effects, which induces a ceiling on power (see SM1.4.1).

One criticism of using the CV to scale the SD of the design prior across effect sizes is that for small treatment effects the required uncertainty becomes very small. However, we argue that this emphasises an important point: when the anticipated effect size is very small, the required prior certainty must be very high to achieve adequate assurance power. For instance, for an expected RD of –1.0% and CV of 0.5, the 95% interval on the design prior is only –2% to 0% (Table SM1.2). For an expected RD<-2.0%, we think the degree of prior certainty that must exist to achieve at least 80% power is unattainable. Consequently, randomised trials targeting very small treatment effects are likely to be substantially underpowered, potentially rendering them unfeasible.

Across the simulations, the sample sizes required to achieve 80% or 90% power were similar for frequentist and Bayesian assurance (Table 1). Differences between the methods are explained by three mechanisms. First, sampling variability. Second, the weakly informative analysis prior used to fit the Bayesian logistic regression models has a mild regularising effect, pulling the posterior mean towards the null value, increasing the required sample size relative to frequentist assurance. Third, the frequentist assurance method involved sampling only from the β_1_ design prior, treating the control event rate as fixed at 50%. By contrast, the Bayesian power method sampled from both β_0_ and β_1_, accommodating uncertainty in both the control event rate and the treatment effect. The variance (SD^2^) of a binomial proportion is maximised at an event rate of 50% and progressively decreases at higher and lower values.^‡,2^ By allowing the control event rate to vary from 50%, the Bayesian method results in lower variance on the pooled proportion, reducing the required sample size for a given power. The aforementioned differences mean the assurance (one-sided P≤0.025) and Bayesian (*P*(*β*_1_ < 0|data) > 0.975) success criteria are similar but not identical. Agreement between the two methods would be improved by assigning flat analysis priors on β_0_ and β_1_; however, we elected not to do this, as weakly informative analysis priors reflect usual practice for Bayesian models.^14^

Normal approximations to Bayesian posterior distributions are commonly used during the design phase of Bayesian randomised trials and are described in statistical texts.^12, 15, 16^ However, the methodology is not widely discussed in the clinical literature. We found almost complete agreement between our normal approximation method and MCMC sampling (see SM1.8, Fig SM1.1). To our knowledge, validation of a normal approximation for to a Bayesian posterior distribution using logistic regression has not been previously reported. The normal approximation allowed us to calculate Bayesian power across a wide range of effect sizes, design priors, and sample sizes and required less than 8 hours of computation time on a modern laptop. Full MCMC sampling would have required days (or weeks) of computation time using MCMC sampling. Consequently, our normal approximation method may be useful for researchers planning Bayesian randomised trials and is freely available to download.

One limitation of the present study is that the simulations were based on an expected event rate of 50%, which is realistic for multicentre trials in critical care targeting interventions for sepsis and acute respiratory distress syndrome.^3^ However, for multicentre trials in anaesthesia, lower event rates are common (e.g., trials investigating surgical site infection or awareness).^4^ At lower event rates, the lower variance reduces the sample size required to achieve a target design power.^2^ While the theoretical power ceiling (Equation 3) is independent of the event rate, the assurance methods described here may become less reliable at very low event rates (<5%) because estimation of the regression coefficients is less robust at extreme event rates.

### Conclusions

Conventional sample size estimates assume a fixed treatment effect. Failure to account for genuine uncertainty in the expected treatment effect means achieved power may be less than design power, which may help explain the low proportion of multicentre trials that report statistically significant differences. Incorporating uncertainty through a design prior may give researchers a more realistic guide to the required sample size and inform decisions on feasibility. We think a CV of around 0.5 is a reasonable default choice for the design prior, as the expected effect size accommodates realistic uncertainty in the effect size but adequate power remains achievable.

## Data Availability

All data produced are included in the manuscript. The R code to replicate the simulations is available at https://github.com/cjdbarlow/papers.

## Supplementary Material

· **Supplementary Material 1.** A detailed explanation of the methods and underlying statistical theory. Supplementary tables. Validation of the normal approximation method to full Markov chain Monte Carlo sampling. Relationship between our frequentist assurance method and that of Markus Huber. Derivation of the ceiling power. Relationship to the ceiling power derived by De Santis and Gubbiotti.
· **Supplementary Material 2**. Full table showing power at different sample size and the power required to achieve 80% and 90% power across effect sizes, design priors, and sample sizes.

## Declaration of interests

DS is an associate editor and member of the editorial board for BJA Education. JB does not have any conflicts of interests to declare.

## Declaration on generative AI and AI-assisted technologies in the manuscript preparation process

During the preparation of this work we used Claude (Anthropic) to draft the code in R to do the simulations and to generate the figures and tables. After using this service, we reviewed and edited the code and carefully evaluated the methodology, tables, and figures. We take full responsibility for the content of the published article.

## Funding

We received no funding for this study.

## Data sharing

The R code used in all the simulations is available at https://github.com/cjdbarlow/papers.

## Author contributions

DS conceived the project, planned the analysis, analysed data, wrote the first draft, edited the manuscript, approved the final version. JB edited the R code, analysed the data, edited the manuscript, approved the final version.

## Footnotes

* Analysis priors are distinct from design priors. Design priors are used to represent uncertainty in the expected event rates and analysis priors are used for fitting the Bayesian logistic regression models.

† 95% distribution for the expected effect size. Because the CV is defined on the log scale, the distribution on the probability scale is not symmetrical.

‡ Variance for a single binomial proportion is *σ*^2^ = *p*(1 − *p*), where *p* is the success probability. The expression is maximised when *p* = 0.5 and progressively decreases for *p* > 0.5 and *p* < 0.5.

