## Supplementary Material 1 for "Accounting for uncertainty in the expected treatment effect substantially increases the sample size required for randomised trials: implications for the feasibility of clinical trials in anaesthesia and critical care"

#### Glossary and notation

| | |
|---|---|
| $\Phi(.)$ | Cumulative distribution function (c.d.f) for the standard normal distribution. |
| $q_{1-\alpha}$ | The (1-α)-quantile of the standard normal distribution. The quantile function is the inverse of the c.d.f function: $q_{1-\alpha} = \Phi^{-1}(1-\alpha)$. |
| $N(\mu, \sigma^2)$ | Normal distribution with mean $\mu$ and standard deviation $\sigma$. |
| Standardisation | Converting $X \sim N(\mu, \sigma^2)$ to the standard normal distribution, $Z \sim N(0,1)$. For any random variable $X$, $P(X \leq x) = P\left(\frac{X-\mu}{\sigma} < \frac{x-\mu}{\sigma}\right) = P\left(Z < \frac{x-\mu}{\sigma}\right) = \Phi\left(\frac{x-\mu}{\sigma}\right)$. If we are calculating $P(X \leq 0)$, we have $P\left(Z < \frac{0-\mu}{\sigma}\right) = \Phi\left(\frac{-\mu}{\sigma}\right)$. |
| $\beta_0$, $\beta_1$ | Population values for the logistic regression coefficients. $\beta_0$ is the intercept coefficient and represents the log odds of the event rate in the control group. $\beta_1$ is the slope coefficient and represents the log odds ratio of the intervention group relative to the control group. |
| $\hat{\beta}_0, \hat{\beta}_1$ | Observed values for the logistic regression coefficients from fitted model. |
| $E(.)$ | The expected (mean) value. |
| $\eta_i$ | The linear predictor for a logistic regression model; i.e. $\beta_0 + \beta_1 z_i$. |
| $\text{logit}(p_i)$ | The logit function, where $\text{logit}(p_i) = \log\left(\frac{p_i}{1-p_i}\right)$ where log is the natural logarithm. The logit function is the inverse of the logistic function, so $\text{logit}(p_i) = \eta_i$. |
| $\text{logistic}(\eta_i)$ | The logistic function: $\text{logistic}(\eta_i) = \frac{\exp(\eta_i)}{1+\exp(\eta_i)}$, where exp is the natural exponential function. The logistic function is the inverse of the logit function, so $\text{logistic}(\eta_i) = p_i$. |
| Design priors | Used to represent uncertainty in the expected baseline event rate ($\beta_0$) and treatment effect ( $\beta_1$). For all simulations, we used an informative prior on $\beta_0$, $N(0, 0.1^2)$. We varied the design prior on $\beta_1$ based on effect size and coefficient of variation but all were informative. |
| Analysis prior | Used for fitting the Bayesian logistic regression models for Bayesian power. For $\beta_0$ we used $N(0, 0.5^2)$ for all simulations, which is centred on an event rate of 0.5 and weakly informative. For $\beta_1$, we used $N(0, 0.5^2)$, which is neutral (centred on the null effect) and weakly informative. |

**SM.1 The statistical model**

The statistical model is logistic regression with a binary response variable $Y$ (event versus no event) and a binary explanatory variable (group allocation: control versus intervention), denoted $z$. We define:

$$Y_i = \begin{cases} 1, & \text{event} \\ 0, & \text{no event} \end{cases}$$

$$z_i = \begin{cases} 1, & \text{intervention} \\ 0, & \text{control} \end{cases}$$

For the $i^{th}$ patient, $Y_i$ is assumed to follow a Bernoulli distribution $Y_i \sim$ Bernoulli$(p_i)$, where $p_i$ is a probability and $E(Y_i) = p_i$. The link is the logit function. So, the model is:

$$\text{logit}(p_i) = \beta_0 + \beta_1 z_i, \qquad \text{(SM.1)}$$

where $\beta_0$ is the intercept coefficient, and $\beta_1$ is the slope coefficient. $\beta_0$ represents the log odds of the event in the control group and $\beta_1$ represents the log odds ratio (OR) of the event in the intervention group relative to the control group. The expression $\beta_0 + \beta_1 z_i$ is the linear predictor, denoted $\eta$. And:

- $\beta_1 > 0$ implies the treatment is harmful.
- $\beta_1 < 0$ implies the intervention is beneficial.

The event probabilities in the control ($p_C$) and intervention($p_I$) groups are given by:

$$p_C = \text{logistic}(\beta_0) \qquad \text{(SM.2)}$$

$$p_I = \text{logistic}(\beta_0 + \beta_1) \qquad \text{(SM.3)}$$

**SM.2 Power functions for conventional frequentist power and frequentist assurance**

Throughout the paper, we calculate the conventional frequentist power (using fixed values for the event rates) and the frequentist assurance power (using a random draw from $\beta_1$, see below) using the power function for a two-armed trial with a binary outcome and two equal sized groups with a one-sided test of $\alpha = 0.025$.

*SM.2.1 Conventional power function*

For a one-sided test at significance threshold $\alpha$:

$$\text{power} = \Phi\left(\frac{|p_C - p_I|}{SE} - q_{1-\alpha}\right), \qquad \text{(SM.4)}$$

where $SE$ is the standard error of the difference in sample proportions:

$$SE = \sqrt{\frac{p_C(1-p_C)}{n_g} + \frac{p_I(1-p_I)}{n_g}}, \qquad \text{(SM.5)}$$

where, $n_g$ is the group size. For all frequentist calculations, we assume $p_C = 0.5$ and $\alpha = 0.025$, where $q_{1-\alpha} = q_{0.975} = 1.96$. So, Equations SM.4 and SM.5 become:

$$\text{power} = \Phi\left(\frac{|0.5 - p_I|}{SE} - 1.96\right) \qquad \text{(SM.6)}$$

and

$$SE = \sqrt{\frac{0.25}{n_g} + \frac{p_I(1-p_I)}{n_g}}. \qquad \text{(SM.7)}$$

Equation SM.6 is the power function that we used for the conventional frequentist power curves that appear on all plots.

*SM.2.2 Assurance power function*

We now derive the equations for the frequentist assurance power curves using the logistic regression model. First, note when $p_C = 0.5$, Equation SM.3 becomes:

$$p_I = \text{logistic}(\beta_0 + \beta_1) = \text{logistic}(\text{logit}(0.5) + \beta_1) = \text{logistic}(\beta_1) \quad \text{(SM.8)}^{*}$$

Mapping Equations SM.6 and SM.7 to the logistic regression model, we have:

$$\text{power} = \Phi\left(\frac{|0.5 - \text{logistic}(\beta_1)|}{SE} - 1.96\right) \quad \text{and} \quad \text{(SM.9)}$$

$$SE = \sqrt{\frac{0.25}{n_g} + \frac{\text{logistic}(\beta_1)(1 - \text{logistic}(\beta_1))}{n_g}}. \quad \text{(SM.10)}$$

Equation SM.9 is the power function used for calculating assurance power, as described below.

**SM.3 Design priors**

The design priors reflect our uncertainty in the true effect size. For the logistic regression model, each design prior involves placing a prior on $\beta_0$, representing the log odds of the event rate in the control group, and a prior on $\beta_1$, representing the log OR of the event in the intervention group relative to the control group.

*SM.3.1 Design prior on* $\beta_0$

For every simulation, we used:

---

[*] Since $\text{logit}(0.5) = \log(0.5/1 - 0.5) = \log(1) = 0$.

$$f(\beta_0) \sim N(0, 0.1^2) \quad \text{(SM.11)}$$

On the log odds scale, a mean of 0 represents odds of 1 ($\exp(0) = 1$). Since, $0.5/(1 - 0.5) = 1$, odds of 1 equates to an event probability of 0.5. The standard deviation (SD) on the log odds scale corresponds to 95% of the probability density between an event rate of 0.45 and 0.55. Thus, the prior on $\beta_0$ is strongly informative.

*SM.3.2 Design priors on $\beta_1$*

At each sample size, we used 10 different normal priors on $\beta_1$ representing different strengths of belief in the treatment effect. To scale the SD across the different effect sizes, we fixed the coefficient of variation (CV). We defined CV as:

$$CV = \frac{\sigma}{|\mu|}, \quad \text{(SM.12)}$$

where $\sigma$ represents the SD and $|\mu|$ represents the absolute value of the mean. By scaling the SD to the CV, the probability mass assigned to a beneficial treatment effect (i.e., $P(\beta_1 < 0)$ or equivalently $P(\log(OR) < 0)$ or $P(\mathrm{OR} < 1)$) was constant across all effect sizes. Table SM.1 shows the prior probability of benefit at each of the CV levels.

**Table SM.1** Design priors on $\beta_1$ based on coefficient of variation (CV).

| Coefficient of variation | Prior probability of benefit ($P(\beta_1 < 0)$) | Clinical interpretation |
|---|---|---|
| 0.1 | ≈100% | Extreme confidence |
| 0.2 | ≈100% | |
| 0.3 | ≈100% | |
| 0.4 | 99% | |
| 0.5 | 98% | High confidence |
| 0.75 | 91% | |
| 1.0 | 84% | Moderate confidence |
| 1.19 | 80% | |
| 1.5 | 75% | |
| 2.0 | 69% | |

Table SM.2 shows the 95% intervals for the risk difference (RD) for each of the design priors on $\beta_1$ across effect sizes.

**Table SM.2.** 95% intervals for the RD for the design priors by coefficient of variation across the different effect sizes. † denotes intervals that cross zero.

| Expected RD -0.10 (-10%) (OR=0.67, $\beta_1$=-0.41) | |
|---|---|
| Coefficient of variation | 95% interval for the RD |
| 0.10 | (-0.119, -0.081) |
| 0.20 | (-0.137, -0.061) |
| 0.30 | (-0.156, -0.042) |
| 0.40 | (-0.173, -0.022) |
| 0.50 | (-0.191, -0.002) |
| 0.75 | (-0.231, +0.048) † |
| 1.00 | (-0.269, +0.096)† |
| 1.19 | (-0.294, +0.132)† |
| 1.50 | (-0.332, +0.187)† |
| 2.00 | (-0.380, +0.266)† |

| Expected RD -0.05 (-5%) (OR=0.82, $\beta_1$=-0.20) | |
|---|---|
| Coefficient of variation | 95% interval for the RD |
| 0.10 | (-0.060, -0.040) |
| 0.20 | (-0.069, -0.030) |
| 0.30 | (-0.079, -0.021) |
| 0.40 | (-0.089, -0.011) |
| 0.50 | (-0.098, -0.001) |
| 0.75 | (-0.121, +0.024)† |
| 1.00 | (-0.144, +0.048)† |
| 1.19 | (-0.161, +0.066)† |
| 1.50 | (-0.188, +0.096)† |
| 2.00 | (-0.229, +0.142)† |

| Expected RD of -0.02 (-2%) (OR=0.92, $\beta_1$=-0.08) | |
|---|---|
| Coefficient of variation | 95% interval for the RD |
| 0.10 | (-0.024, -0.016) |
| 0.20 | (-0.028, -0.012) |
| 0.30 | (-0.032, -0.008) |
| 0.40 | (-0.036, -0.004) |
| 0.50 | (-0.040, -0.000) |
| 0.75 | (-0.049, +0.009)† |
| 1.00 | (-0.059, +0.019)† |
| 1.19 | (-0.066, +0.027)† |
| 1.50 | (-0.078, +0.039)† |
| 2.00 | (-0.097, +0.058)† |

| Expected RD of -0.01 (-1%) (OR=0.96, $\beta_1$=-0.04) | |
|---|---|
| Coefficient of variation | 95% interval for the RD |
| 0.10 | (-0.012, -0.008) |
| 0.20 | (-0.014, -0.006) |
| 0.30 | (-0.016, -0.004) |
| 0.40 | (-0.018, -0.002) |
| 0.50 | (-0.020, -0.000) |
| 0.75 | (-0.025, +0.005)† |
| 1.00 | (-0.030, +0.010)† |
| 1.19 | (-0.033, +0.013)† |
| 1.50 | (-0.039, +0.019)† |
| 2.00 | (-0.049, +0.029)† |

Notice that by applying a constant CV, the design priors for $\beta_1$ impose very tight intervals on the range of treatment effects when the expected effect size is very small. This might be interpreted as a criticism of using the CV to scale the SD across the effect sizes. However, imposing a constant CV maintains a constant degree of uncertainty across the effect sizes. An alternative interpretation is that for small effect sizes, high prior certainty is required to achieve adequate power, which itself is an important in terms of the feasibility of trials targeting small effects.

**SM.4 Frequentist assurance**

The frequentist success criterion was a one-sided p-value ≤0.025. Assurance power is obtained by integrating the power function over the design prior:

$$\text{Assurance power} = \int_{-\infty}^{0} \text{power}(\beta_1, n_g) f(\beta_1) d\beta_1, \quad \text{(SM.13)}$$

where $\text{power}(\beta_1, n_g)$ is the power function given in Equation SM.9 and $f(\beta_1)$ is the design prior on $\beta_1$. Since, only negative values for $\beta_1$ contribute to power (one-sided test), the limits of integration are 0 to $-\infty$.

The integral in Equation SM.13 has no closed form and we computed assurance power using Monte Carlo integration, whereby we averaged the power across 5,000 random samples drawn from the design prior on $\beta_1$. Draws where $\beta_1 \geq 0$, representing neutral or harmful effects, were assigned power of 0, to ensure directional consistency with the one-sided significance level of 0.025. We computed the assurance power for each effect size, CV level, and sample size. The sample sizes and steps for the assurance power calculations are shown in Table SM.3.

**Table SM.3**. Frequentist assurance sample sizes, step sizes, and number of draws from $\beta_1$.

| Scenario | Sample size (step size) | N steps | CV levels | Draws per CV and steps | Total draws |
|---|---|---|---|---|---|
| RD = −10% | 200-4,000 (200)<br>4,500-10,000 (500) | 37 | 10 | 5,000 | 1,850,000 |
| RD = −5% | 200-4,000 (200)<br>5,000-20,000 (1,000) | 36 | 10 | 5,000 | 1,800,000 |
| RD = −2% | 500-5,000 (500)<br>7,500-50,000 (2,500) | 28 | 10 | 5,000 | 1,400,000 |
| RD = −1% | 1,000-10,000 (1,000)<br>15,000-100,000 (5,000) | 28 | 10 | 5,000 | 1,400,000 |

*SM.4.1 Comparison to the method described by Markus Huber*

In a recent correspondence Markus Huber provided an important illustration of frequentist assurance applied to a dataset of multicentre randomised trials in anaesthesia and critical care, including trials previously studied by our group.[1, 2] To our knowledge, Huber's paper is the first application of assurance in a clinical anaesthesia journal. Our paper builds on this contribution by extending the framework in two ways. First, Huber used a uniform design prior bounded between zero effect and the assumed effect size for each trial. The uniform prior has the advantage of simplicity but places no probability mass on harmful effects and does not reflect a coherent prior belief about the treatment effect. By contrast, our normal design prior is more flexible, allows systematic exploration of uncertainty across a range of beliefs, and assigns some probability mass to harmful effects, which is clinically realistic. Second, Huber's prior always assigns a probability of 1 to the alternative hypothesis, meaning the theoretical ceiling on power is always 100%. Our normal design prior parameterised by CV makes the ceiling explicit and clinically interpretable (see below).

**SM.5 Bayesian power**

The Bayesian success criterion was $P(OR < 1|\text{data}) > 0.975$, which can also be expressed as $P(\log(OR) < 0|\text{data}) > 0.975$ or $P(\beta_1 < 0|\text{data}) > 0.975$. The Bayesian success criterion is analogous to the frequentist criterion of a one-sided p-value ≤0.025.

Datasets were simulated as follows. For each simulation at a particular sample size ($n_i$), we sampled one value each from the design priors on $\beta_0$ and $\beta_1$ and converted the values to event probabilities for the control and intervention groups ($p_C$, $p_I$) using Equations SM.2 and SM.3. For each simulation, we generated $n_i$ outcomes by drawing $n_i/2$ random samples from two Binomial distributions:

$X_C \sim \text{Binomial}(n_i/2\,, p_C)$

$X_I \sim \text{Binomial}(n_i/2\,, p_I)$

The sizes and steps for the simulated datasets for Bayesian power are shown in Table SM.4.

**Table SM.4.** Simulated datasets for Bayesian power.

| Scenario | Sample size (step size) | N steps | CV levels | Simulated trials per CV and step | Total trials |
|---|---|---|---|---|---|
| RD = −10% | 200-4,000 (200)<br>4,500-10,000 (500) | 37 | 10 | 2,000 | 740,000 |
| RD = −5% | 200-4,000 (200)<br>5,000-20,000 (1,000) | 36 | 10 | 2,000 | 720,000 |
| RD = −2% | 500-5,000 (500)<br>7,500–50,000 (2,500) | 28 | 10 | 2,000 | 560,000 |
| RD = −1% | 1,000-10,000 (1,000)<br>15,000-100,000 (5,000) | 28 | 10 | 1,000 | 280,000 |

For each simulated dataset we fitted a Bayesian logistic regression model using analysis priors. For each model we specified identical analysis priors on $\beta_0$ and $\beta_1$:

$$f(\beta_0) = f(\beta_1) \sim N(0, 0.5^2) \quad \text{(SM.14)}$$

For $\beta_0$, the analysis prior is centred on a log odds of 0 (odds of 1), which is equivalent to an expected control event rate of 0.5 with 95% coverage of 0.27 to 0.72 on the probability scale. For $\beta_1$, the analysis prior is centred on a log OR of 0 (OR of 1), which is equivalent to an expected RD of 0, with 95% coverage of 0.38 to 2.67 on the OR scale and -0.23 to 0.23 on the RD scale. Thus, the analysis prior on $\beta_1$ is neutral and weakly informative and the analysis prior on $\beta_0$ is centred on the expected event rate and weakly informative.

We estimated posterior distributions using the normal approximation method, described below. The normal approximation method was verified against full Markov chain Monte Carlo (MCMC) sampling for a limited number of power curves (see below). From each posterior distribution, we extracted the posterior mean for $\beta_1$ and determined whether it met the success criterion. The proportion of posterior distributions at each sample size that met the success criterion was defined as the Bayesian power for that effect size and CV.

**SM.6 Equivalence with two-sided tests at the 5% threshold**

When the design prior is centred on a beneficial treatment effect (OR<1, RD<0) the frequentist assurance ($P \leq 0.025$, one-sided test) and Bayesian success criteria ($P(\beta_1 < 0|\text{data}) > 0.975$) used in the simulations are equivalent to two-sided tests at the 5% threshold. We can write the two-sided success criteria for assurance as $P \leq 0.05$ (two-sided test) and for Bayesian power as $P(\beta_1 < 0|\text{data}) > 0.975$ or $P(\beta_1 > 0|\text{data}) > 0.975$. When the design prior has most of its probability mass in the region of benefit (our choices range from approximately 100% (CV of 0.1) to 69% (CV of 2.0), the upper tail success criterion (i.e. detecting harm with >0.975 probability) is never satisfied and contributes 0 power. The equivalence breaks down if the design prior were centred on the null effect or had a substantial part of its probability mass in the region of harm.

**SM.7 Frequentist assurance versus Bayesian power: similar but not equivalent**

The frequentist assurance ($P \leq 0.025$) and Bayesian ($P(\beta_1 < 0|\text{data}) > 0.975$) success criteria are similar but not equivalent. First, when fitting the Bayesian logistic regression model to obtain the posterior distribution, we specified weakly informative analysis priors for $\beta_0$ and $\beta_1$ (Equation SM.14). The weakly informative analysis priors have a mild regularising effect, pulling the posterior mean towards to the null value, increasing the required sample size to satisfy the success criterion. Second, for the frequentist assurance method, we only sampled from the design prior on $\beta_1$ and treated the control event rate as fixed at 0.5, whereas for Bayesian power we sampled from $\beta_0$ and $\beta_1$. By allowing the control event rate to vary from 0.5, the Bayesian method results in lower variance on the difference in proportions, reducing the required sample size required to satisfy the success criterion. With these two opposing effects, differences between the two methods were small (see Figures 1-4, main manuscript).

**SM.8 The normal approximation to the posterior**

For the normal approximation method we make two assumptions:

- *Assumption 1*. That the joint posterior distribution $f(\beta_0, \beta_1|\text{data})$ approximates a bivariate normal distribution.[3] Consequently, the marginal posterior distribution for $\beta_1$ ($f(\beta_1|\text{data})$) is also approximately normal.
- *Assumption 2*. That the sampling distribution for $\hat{\beta}_1$is approximately normally distributed, with mean $\beta_1$ and variance $SE(\beta_1)$.[4] That is,

$$\hat{\beta}_1 \approx N(\beta_1, SE(\beta_1)^2). \qquad \text{(SM.15)}$$

Substituting the maximum likelihood estimate and SE for $\beta_1$ from the fitted logistic regression model, we have:

$$\hat{\beta}_1 \approx N(\hat{\beta}_1, \widehat{SE}^2). \qquad \text{(SM.16)}$$

Denoting the analysis prior on $\beta_1$ as $N(a, b)$, $a$ is the mean and $b$ is the variance, we then applied the normal-normal conjugate model. For the normal-normal conjugate model with known variance, the closed form posterior distribution for a single observation on $\beta_1$ is:

$$\beta_1|\text{data} \sim N\left(\frac{\widehat{SE}^2 a + \hat{\beta}_1 b}{\widehat{SE}^2 + b}, \frac{\widehat{SE}^2 b}{\widehat{SE}^2 + b}\right). \qquad \text{(SM.17)}$$

Denoting the posterior mean as $\mu_{\text{post}}$ and variance as $\sigma^2_{\text{post}}$, we can rearrange the terms in Equation SM.17 to obtain:

$$\sigma_{\text{post}}^2 = \frac{1}{\frac{1}{b}+\frac{1}{\widehat{SE}^2}} \quad \text{and} \qquad ^{\dagger}\text{(SM.18)}$$

$$\mu_{\text{post}} = \sigma_{\text{post}}^2 \left(\frac{a}{b} + \frac{\widehat{\beta}_1}{\widehat{SE}^2}\right). \qquad \text{(SM.19)}$$

As with the full MCMC model, the analysis prior on $\beta_1$ has parameters $a = 0$ and $b = 0.5^2 = 0.25$. So, for our simulations, we have $\sigma_{\text{post}}^2 = \frac{1}{4+\frac{1}{\widehat{SE}^2}}$ and $\mu_{\text{post}} = \sigma_{\text{post}}^2 \left(\frac{\widehat{\beta}_1}{\widehat{SE}^2}\right)$.

Standardising (see Glossary), we can write the success criterion as:

$$P(\beta_1 < 0|\text{data}) = \Phi\left(\frac{0-\mu_{\text{post}}}{\sigma_{\text{post}}}\right) \quad \text{or}$$

$$P(\beta_1 < 0|\text{data}) = \Phi\left(\frac{-\mu_{\text{post}}}{\sigma_{\text{post}}}\right) \qquad \text{(SM.20)}$$

*SM.8.1 What about the analysis prior on $\beta_0$?*

The analysis prior on $\beta_0$ is not explicitly incorporated in the normal-normal conjugate model, which requires justification. In a full Bayesian model, $\beta_0$ and $\beta_1$ are estimated jointly and their posterior distributions are correlated – meaning that the prior on $\beta_0$ can, in principle, influence the posterior for $\beta_1$. However, for a balanced two-arm trial, the correlation between the two parameters is small, and the indirect influence of the $\beta_0$ prior on the posterior for $\beta_1$ is modest. Furthermore, when the analysis prior on $\beta_0$ is centred on the observed event rate in the control group, as in the present application, it contributes

---

† Equivalently, using precision instead of variance (where precision is the reciprocal of variance) the posterior precision is $\frac{1}{b} + \frac{1}{\widehat{SE}^2}$. Precision was used in the R code.

little information beyond what the likelihood already provides, further reducing its influence. Finally, as we demonstrate in S2, there was excellent agreement between MCMC sampling and the normal approximation.

*SM.8.2 Steps in the normal approximation*

Summarising the steps in the normal approximation, we have:

1. Generate simulated data at sample size $n_i$ from the design priors for $\beta_0$ and $\beta_1$.

2. Fit a standard logistic regression model by maximum likelihood.

3. Extract the maximum likelihood estimate for the $\beta_1$ coefficient (i.e., $\hat{\beta}_1$ from Equation SM.16) and standard error ($SE(\hat{\beta}_1)$, also from Equation SM.16) from the *fitted logistic regression model*. Note this $\beta_1$ coefficient is from the fitted model – not the design prior.

4. Calculate the closed form of the posterior mean using the analysis prior on $\beta_1$ (parameters $a = 0$ and $b = 0.5^2 = 0.25$.) and the normal-normal conjugate model (Equations SM.18 and SM.19).

5. Calculate the posterior probability of success and determine if it satisfies the success criterion.

6. Repeat steps 1-5 for $N$ simulations and calculate Bayesian power as the proportion of simulations in which the success criterion is satisfied

Note, when calculating the normal approximation, there are three distinct forms of the $\beta_1$ coefficient: (1) the $\beta_1$ coefficient from the design prior that is used for data generation to fit the logistic regression model; (2) the $\hat{\beta}_1$ coefficient estimated by maximum likelihood from the regression fitted model; (3) the $\beta_1$ coefficient for the analysis prior (with mean 0 and variance $0.5^2$) used for the conjugate update.

**SM.9 Full MCMC sampling for Bayesian power: validation with the normal approximation**

For validation of the normal approximation method, we fitted Bayesian logistic regression models in R using the brms package.[5] The brms package uses RStan as the interface between R and the probabilistic programming language Stan.[6] Stan implements the No-U-Turn Sampler (NUTS) for MCMC sampling, which is a modification of Hamiltonian Monte Carlo.[7] Full MCMC sampling for calculating the Bayesian power across the simulations would have taken weeks of computation time. However, we validated the normal approximation against full MCMC sampling using a limited range of simulations.

Figure SM.1 shows the results obtained from full MCMC sampling and the normal approximation method across effect sizes for a selection of CV values (0.1, 0.5, 1.19, 1.5). To reduce the computation time, we reduced the number of simulations per step to 1,000 (from 2,000). We can see from Figure SM.1 that there is very close agreement between the two methods across effect sizes, CVs, and sample sizes.

**Figure SM.1**. Validation of the normal approximation method with full MCMC sampling.

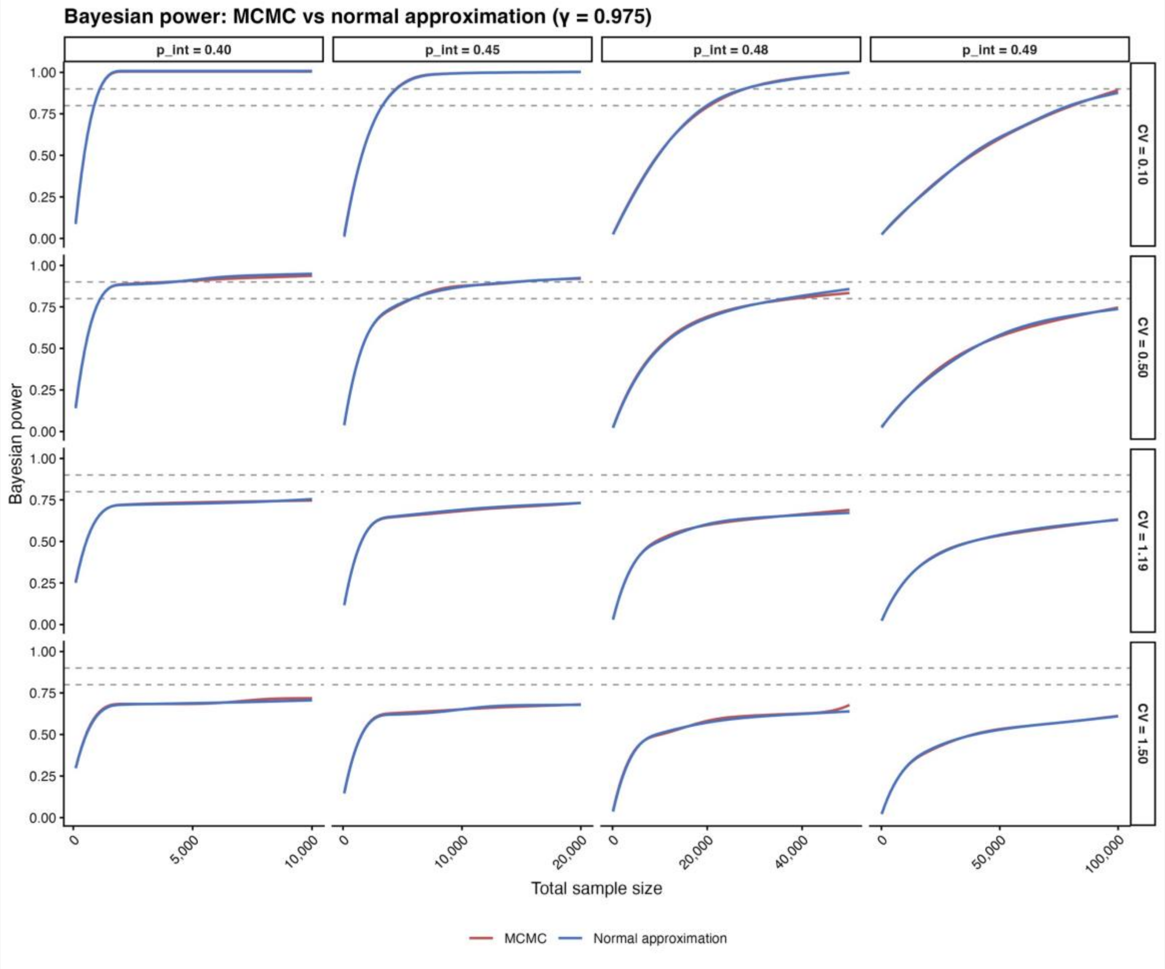


### SM.10 Theoretical power limit

In this section, we derive the theoretical upper limit on achieved power as the sample size increases without bound (i.e. $n \to \infty$), which we denote $\mathrm{power}_\infty$. We have the Bayesian and frequentist success criteria:

- *Bayesian*: $P(\beta_1 < 0|\mathrm{data}) > 0.975$. That is, the posterior probability of a beneficial treatment effect exceeds 0.975.

- *Frequentist*: $p \leq 0.025$ where $p = 1 - \Phi(z)$ for $\hat{z} = \frac{-\hat{\beta}_1}{SE(\beta_1)}$ where $Z \sim N(0,1)$ under the null hypothesis ($H_0: \beta_1 \geq 0$, $H_1: \beta_1 < 0$). That is, a one-sided p-value less than or equal to 0.025.

Now consider what happens as $n \to \infty$. For the Bayesian model, the posterior SD falls to 0 and the posterior distribution concentrates at a point mass at $\beta_1$. For the frequentist model, the SE falls to zero and the z-statistic increases without bound to either $-\infty$ (when $\hat{\beta}$ is positive) or $\infty$ (when $\hat{\beta}$ is negative). For a single random draw from the design prior as $n \to \infty$, we have three possibilities:

1. The true effect is harmful ($\beta_1 > 0$)
   a. *Bayesian*. As $n \to \infty$, the posterior distribution ($\beta_1|\text{data}$) is concentrated above 0 and the Bayesian success criterion is never satisfied.
   b. *Frequentist*. As $n \to \infty$, $\hat{\beta}_1 \to \beta_1 > 0$ and the $SE \to 0$, so $z \to -\infty$. When $z \to -\infty$ the p value tends to 1 (since $p = 1 - \Phi(-\infty) = 1 - 0 = 1$). So, the frequentist success criterion is never satisfied.
2. The true effect is zero ($\beta_1 = 0$)[‡]
   a. *Bayesian*. As $n \to \infty$, the posterior distribution ($\beta_1|\text{data}$) is concentrated at 0 and the Bayesian success criterion is never satisfied.

---

[‡] Note, this scenario requires that the true effect is exactly zero – which is impossible for a continuous variable (a measure-zero event); so in practice, events where $\beta_1 = 0$ do not contribute to the power calculations.

b. *Frequentist*. As $n \to \infty$, $\hat{\beta}_1 \to \beta_1 = 0$ and $SE \to 0$, so $z \to 0$. When $z \to 0$, the p-value tends to 0.5 (since (since $p = 1 - \Phi(0) = 1 - 0.5 = 0.5$), so the success criterion is never satisfied.

3. True effect is beneficial ($\beta_1 < 0$)

   a. *Bayesian*. As $n \to \infty$, the posterior distribution ($\beta_1|\text{data}$) is concentrated below 0 and the Bayesian success criterion is always satisfied.

   b. *Frequentist*. As $n \to \infty$, $\hat{\beta}_1 \to \beta_1 < 0$ and $SE \to 0$, so $z \to \infty$. When $z \to \infty$ the p value tends to 0 (since $p = 1 - \Phi(\infty) = 1 - 1 = 0$). So, the frequentist success criterion is always satisfied.

So, in the limit, as $n \to \infty$, both the frequentist and Bayesian success criteria are satisfied if $\beta_1 < 0$. Defining the upper bound on the power as $\text{power}_\infty$ (i.e. $\text{power}_\infty = \lim_{n\to\infty} \text{Power}$), we can write

$$\text{power}_\infty = P(\beta_1 < 0) \qquad \text{(SM.21)}$$

The design prior on $\beta_1$ is normal with $N(\mu, \sigma^2)$ where $\mu < 0$ and $\sigma > 0$. Standardising (see Glossary) we have:

$$P(\beta_1 < 0) = P\left(\frac{\beta_1 - \mu}{\sigma} < \frac{0-\mu}{\sigma}\right) = P\left(Z < \frac{-\mu}{\sigma}\right), \qquad \text{where } Z \sim N(0,1).$$

So,

$$P(\beta_1 < 0) = P\left(Z < \frac{-\mu}{\sigma}\right) = \Phi\left(\frac{-\mu}{\sigma}\right).$$

Since $\mu < 0$, then $\frac{-\mu}{\sigma} = \frac{|\mu|}{\sigma} = \frac{1}{CV}$, where CV is the coefficient of variation for the design prior, defined as $CV = \frac{\sigma}{|\mu|}$. So,

$$\text{power}_\infty = \Phi\left(\frac{-\mu}{\sigma}\right) = \Phi\left(\frac{1}{CV}\right). \quad \text{(SM.22)}$$

Notice that the upper bound on power ($\text{power}_\infty$), depends only on the CV of the design prior and holds true for both the Bayesian and frequentist decision criteria and is standardised to the effect size.

Notice also that as $n \to \infty$, the success criterion reduces to $P(\beta_1 < 0)$ (Equation SM.21). Thus, $\text{power}_\infty$ is identical to the probability of a beneficial treatment effect for the design prior. Consequently, the values for $\text{power}_\infty$ are the same as the prior probability of benefit shown in Table SM.1.

**SM.11 Comparison to De Santis and Gubbiotti limit theorem**

De Santis and Gubbiotti demonstrated that for the hypothesis structure $H_0: \theta \leq \theta_0$ and $H_1: \theta > \theta_0$, as $n \to \infty$, $\text{power}_\infty = P(\theta > \theta_0)$ (their case A).[8] Assuming $\theta_0 = 0$, we can write: $\text{power}_\infty = P(\theta > 0)$, which is analogous to our result SM.21.

De Santis and Gubbiotti assume normally distributed data $X \sim N(\theta, \sigma^2)$ where the variance ($\sigma^2$) is known. They summarise the data by the sample mean ($\bar{X}_n$), where, by the central limit theorem:

$$\bar{X}_n \approx N\left(\theta, \frac{\sigma^2}{n}\right) \quad \text{(SM.23)}$$

They specify a normal design prior $\theta \sim N(\theta_d, \sigma^2/n_d)$, where $\theta_d$ is the prior mean and $n_d$ is a prior sample size parameter that scales the prior variance relative to $\sigma^2$. So, the prior SD is

$\sigma_d = \sigma/\sqrt{n_d}$. The reason to include the parameter $n_d$ in the prior precision (reciprocal of variance) is to map to the precision of the sample mean (Equation SM.23.).[§]

Using the normal-normal conjugate model De Santis and Gubbiotti derive their limit theorem as $\text{power}_\infty = 1 - \Phi\left(\frac{-\theta_d\sqrt{n_d}}{\sigma}\right)$, which by the symmetry of the standard normal distribution can be written as:

$$\text{power}_\infty = P(\theta > 0) = \Phi\left(\frac{\theta_d\sqrt{n_d}}{\sigma}\right). \tag{SM.24}$$

With our frequentist assurance approach, we are considering the hypothesis structure $H_0\colon \beta_1 \geq 0$ and $H_1\colon \beta_1 < 0$. The design prior on $\beta_1$ has mean $\mu$ and SD $\sigma$. Since, we are considering the mean on the log OR scale (which is centred on 0), when calculating CV we must use the absolute value of the mean (i.e., $|\mu|$). Table SM.5 compares the parameters of the respective approaches. We can see immediately see that Equations SM.24 and SM.22 are equivalent.

---

[§] In the normal-normal model used by De Santis, they were working with the sample mean as the statistic. In their case, the posterior precision is obtained by adding the prior precision and the likelihood precision: $\frac{1}{\sigma^2_{\text{post}}} = \frac{1}{\sigma^2/n_d} + \frac{1}{\sigma^2/n} = \frac{n_d+n}{\sigma^2}$. The prior contributes $n_d$ units of precision and the data contributes $n$ units of precision. The posterior mean is a weighted average of the prior mean and the data mean, with weights proportional to $n_d$ and $n$, $\mu_{\text{post}} = \frac{n_d\theta_d+n\bar{X}}{n_d+n}$. So, if $n_d = n$, the prior and the data contribute equally to the posterior mean. With our normal approximation method we are working directly with $\hat{\beta}_1$ as the statistic, obtained from the fitted logistic regression model (Equation SM.16), which already has its own standard error (i.e., accounts for the sample size), so there was no need to consider $n_d$.

**Table SM.5.** Comparison of parameters

| Parameter | De Santis and Gubbiotti | Our formulation |
|---|---|---|
| Design prior mean | $\theta_d$ | $\|\mu\|$ |
| Design prior SD | $\sigma/\sqrt{n_d}$ | $\sigma$ |
