## Supplementary Material 2 for "Accounting for uncertainty in the expected treatment effect substantially increases the sample size required for randomised trials: implications for the feasibility of clinical trials in anaesthesia and critical care"

### Risk difference -0.10 (-10%) (OR = 0.67)

Conventional frequentist (α=0.025, one-sided, assumed effect size): N for 80% power = 771 | N for 90% power = 1,083.

***Bayesian assurance power***

| CV | p∞ | Power at n = 300 | Power at n = 800 | Power at n = 6,000 | Power at n = 13,000 | Power at n = 20,000 | N for 80% power | N for 90% power |
|---|---|---|---|---|---|---|---|---|
| 0.10 | 1.00 | 0.336 | 0.776 | 1.000 | 1.000 | 1.000 | 848 | 1,233 |
| 0.20 | 1.00 | 0.353 | 0.747 | 0.999 | 1.000 | 1.000 | 912 | 1,457 |
| 0.30 | 1.00 | 0.367 | 0.713 | 0.987 | 0.997 | 0.997 | 1,000 | 1,803 |
| 0.40 | 0.99 | 0.352 | 0.693 | 0.961 | 0.981 | 0.981 | 1,296 | 1,976 |
| 0.50 | 0.98 | 0.379 | 0.680 | 0.920 | 0.962 | 0.962 | 1,439 | 3,595 |
| 0.75 | 0.91 | 0.391 | 0.641 | 0.830 | 0.852 | 0.879 | 3,216 | - |
| 1.00 | 0.84 | 0.406 | 0.610 | 0.750 | 0.795 | 0.806 | 17,040 | - |
| 1.19 | 0.80 | 0.412 | 0.587 | 0.732 | 0.768 | 0.744 | - | - |
| 1.50 | 0.75 | 0.443 | 0.556 | 0.699 | 0.697 | 0.713 | - | - |
| 2.00 | 0.69 | 0.459 | 0.569 | 0.644 | 0.646 | 0.677 | - | - |

*"-": the target power was not reached within the simulated trial sizes.*

***Frequentist assurance power***

| CV | p∞ | Power at n = 300 | Power at n = 800 | Power at n = 6,000 | Power at n = 13,000 | Power at n = 20,000 | N for 80% power | N for 90% power |
|---|---|---|---|---|---|---|---|---|
| 0.10 | 1.00 | 0.419 | 0.807 | 1.000 | 1.000 | 1.000 | 787 | 1,179 |
| 0.20 | 1.00 | 0.421 | 0.784 | 0.999 | 1.000 | 1.000 | 845 | 1,399 |
| 0.30 | 1.00 | 0.425 | 0.747 | 0.990 | 0.996 | 0.998 | 946 | 1,705 |
| 0.40 | 0.99 | 0.428 | 0.722 | 0.961 | 0.981 | 0.981 | 1,161 | 2,235 |
| 0.50 | 0.98 | 0.437 | 0.694 | 0.926 | 0.951 | 0.953 | 1,530 | 3,509 |
| 0.75 | 0.91 | 0.458 | 0.639 | 0.830 | 0.861 | 0.867 | 3,188 | - |
| 1.00 | 0.84 | 0.454 | 0.619 | 0.772 | 0.788 | 0.807 | 15,426 | - |
| 1.19 | 0.80 | 0.453 | 0.595 | 0.729 | 0.750 | 0.770 | - | - |
| 1.50 | 0.75 | 0.467 | 0.587 | 0.692 | 0.712 | 0.718 | - | - |
| 2.00 | 0.69 | 0.485 | 0.566 | 0.644 | 0.663 | 0.675 | - | - |

*"-": the target power was not reached within the simulated trial sizes.*

### Risk difference -0.05 (-5.0%) (OR = 0.82)

Conventional frequentist (α=0.025, one-sided, assumed effect size): N for 80% power = 3,156 | N for 90% power = 4,232.

***Bayesian assurance power***

| CV | p∞ | Power at n = 400 | Power at n = 900 | Power at n = 9,000 | Power at n = 18,000 | Power at n = 50,000 | N for 80% power | N for 90% power |
|---|---|---|---|---|---|---|---|---|
| 0.10 | 1.00 | 0.124 | 0.307 | 0.993 | 1.000 | 1.000 | 3,361 | 4,491 |
| 0.20 | 1.00 | 0.131 | 0.314 | 0.982 | 0.998 | 1.000 | 3,611 | 4,950 |
| 0.30 | 1.00 | 0.127 | 0.308 | 0.942 | 0.987 | 0.997 | 3,838 | 6,250 |
| 0.40 | 0.99 | 0.150 | 0.324 | 0.894 | 0.950 | 0.980 | 4,531 | 9,667 |
| 0.50 | 0.98 | 0.151 | 0.324 | 0.867 | 0.921 | 0.941 | 5,391 | 14,778 |
| 0.75 | 0.91 | 0.179 | 0.357 | 0.771 | 0.818 | 0.846 | 12,841 | - |
| 1.00 | 0.84 | 0.206 | 0.379 | 0.722 | 0.761 | 0.803 | 49,333 | - |
| 1.19 | 0.80 | 0.233 | 0.401 | 0.665 | 0.723 | 0.750 | - | - |
| 1.50 | 0.75 | 0.249 | 0.426 | 0.674 | 0.677 | 0.715 | - | - |
| 2.00 | 0.69 | 0.306 | 0.441 | 0.605 | 0.634 | 0.633 | - | - |

*"-": the target power was not reached within the simulated trial sizes.*

***Frequentist assurance power***

| CV | p∞ | Power at n = 400 | Power at n = 900 | Power at n = 9,000 | Power at n = 18,000 | Power at n = 50,000 | N for 80% power | N for 90% power |
|---|---|---|---|---|---|---|---|---|
| 0.10 | 1.00 | 0.170 | 0.327 | 0.994 | 1.000 | 1.000 | 3,238 | 4,416 |
| 0.20 | 1.00 | 0.175 | 0.333 | 0.979 | 0.998 | 1.000 | 3,493 | 4,965 |
| 0.30 | 1.00 | 0.177 | 0.337 | 0.947 | 0.982 | 0.995 | 3,820 | 6,218 |
| 0.40 | 0.99 | 0.186 | 0.348 | 0.905 | 0.953 | 0.982 | 4,585 | 8,758 |
| 0.50 | 0.98 | 0.197 | 0.362 | 0.860 | 0.914 | 0.947 | 5,729 | 14,113 |
| 0.75 | 0.91 | 0.220 | 0.371 | 0.781 | 0.824 | 0.863 | 12,926 | - |
| 1.00 | 0.84 | 0.244 | 0.392 | 0.712 | 0.764 | 0.794 | - | - |
| 1.19 | 0.80 | 0.270 | 0.417 | 0.683 | 0.715 | 0.758 | - | - |
| 1.50 | 0.75 | 0.294 | 0.421 | 0.646 | 0.687 | 0.706 | - | - |
| 2.00 | 0.69 | 0.328 | 0.450 | 0.613 | 0.641 | 0.657 | - | - |

*"-": the target power was not reached within the simulated trial sizes.*

### Risk difference -0.02 (-2.0%) (OR = 0.92)

Conventional frequentist (α=0.025, one-sided, assumed effect size): N for 80% power = 19,617 | N for 90% power = 26,489.

***Bayesian assurance power***

| CV | p∞ | Power at n = 700 | Power at n = 8,000 | Power at n = 40,000 | Power at n = 140,000 | Power at n = 300,000 | N for 80% power | N for 90% power |
|---|---|---|---|---|---|---|---|---|
| 0.10 | 1.00 | 0.060 | 0.432 | 0.976 | 1.000 | 1.000 | 20,736 | 28,177 |
| 0.20 | 1.00 | 0.062 | 0.446 | 0.946 | 0.998 | 1.000 | 21,531 | 32,470 |
| 0.30 | 1.00 | 0.057 | 0.445 | 0.905 | 0.987 | 0.995 | 24,338 | 39,141 |
| 0.40 | 0.99 | 0.073 | 0.420 | 0.853 | 0.961 | 0.980 | 28,544 | 54,271 |
| 0.50 | 0.98 | 0.073 | 0.432 | 0.821 | 0.926 | 0.951 | 36,746 | 90,952 |
| 0.75 | 0.91 | 0.071 | 0.453 | 0.736 | 0.830 | 0.857 | 93,182 | - |
| 1.00 | 0.84 | 0.090 | 0.489 | 0.689 | 0.752 | 0.797 | 286,667 | - |
| 1.19 | 0.80 | 0.096 | 0.487 | 0.685 | 0.732 | 0.755 | - | - |
| 1.50 | 0.75 | 0.108 | 0.477 | 0.641 | 0.693 | 0.718 | - | - |
| 2.00 | 0.69 | 0.153 | 0.485 | 0.596 | 0.644 | 0.662 | - | - |

*"-": the target power was not reached within the simulated trial sizes.*

***Frequentist assurance power***

| CV | p∞ | Power at n = 700 | Power at n = 8,000 | Power at n = 40,000 | Power at n = 140,000 | Power at n = 300,000 | N for 80% power | N for 90% power |
|---|---|---|---|---|---|---|---|---|
| 0.10 | 1.00 | 0.077 | 0.433 | 0.971 | 1.000 | 1.000 | 20,085 | 27,668 |
| 0.20 | 1.00 | 0.078 | 0.439 | 0.946 | 0.999 | 1.000 | 21,913 | 31,346 |
| 0.30 | 1.00 | 0.079 | 0.440 | 0.901 | 0.988 | 0.995 | 24,283 | 39,635 |
| 0.40 | 0.99 | 0.080 | 0.442 | 0.861 | 0.958 | 0.980 | 29,222 | 55,860 |
| 0.50 | 0.98 | 0.082 | 0.442 | 0.821 | 0.922 | 0.948 | 34,534 | 93,724 |
| 0.75 | 0.91 | 0.089 | 0.459 | 0.739 | 0.827 | 0.861 | 77,926 | - |
| 1.00 | 0.84 | 0.100 | 0.461 | 0.688 | 0.771 | 0.793 | 278,939 | - |
| 1.19 | 0.80 | 0.112 | 0.470 | 0.655 | 0.728 | 0.759 | - | - |
| 1.50 | 0.75 | 0.126 | 0.475 | 0.616 | 0.688 | 0.704 | - | - |
| 2.00 | 0.69 | 0.164 | 0.476 | 0.605 | 0.632 | 0.650 | - | - |

*"-": the target power was not reached within the simulated trial sizes.*

### Risk difference -0.01 (-1.0%) (OR = 0.96)

Conventional frequentist (α=0.025, one-sided, assumed effect size): N for 80% power = 78,529 | N for 90% power = 105,359.

***Bayesian assurance power***

| CV | p∞ | Power at n = 700 | Power at n = 8,000 | Power at n = 40,000 | Power at n = 140,000 | Power at n = 300,000 | N for 80% power | N for 90% power |
|---|---|---|---|---|---|---|---|---|
| 0.10 | 1.00 | 0.032 | 0.147 | 0.513 | 0.953 | 0.999 | 80,100 | 108,082 |
| 0.20 | 1.00 | 0.033 | 0.131 | 0.517 | 0.921 | 0.995 | 86,761 | 127,674 |
| 0.30 | 1.00 | 0.030 | 0.137 | 0.506 | 0.879 | 0.970 | 96,556 | 155,510 |
| 0.40 | 0.99 | 0.038 | 0.159 | 0.513 | 0.845 | 0.925 | 115,122 | 239,000 |
| 0.50 | 0.98 | 0.030 | 0.165 | 0.517 | 0.806 | 0.897 | 137,818 | - |
| 0.75 | 0.91 | 0.036 | 0.179 | 0.523 | 0.722 | 0.811 | 294,773 | - |
| 1.00 | 0.84 | 0.049 | 0.213 | 0.519 | 0.673 | 0.734 | - | - |
| 1.19 | 0.80 | 0.046 | 0.231 | 0.502 | 0.672 | 0.706 | - | - |
| 1.50 | 0.75 | 0.053 | 0.247 | 0.520 | 0.619 | 0.659 | - | - |
| 2.00 | 0.69 | 0.059 | 0.310 | 0.493 | 0.608 | 0.635 | - | - |

*“-”: the target power was not reached within the simulated trial sizes.*

***Frequentist assurance power***

| CV | p∞ | Power at n = 700 | Power at n = 8,000 | Power at n = 40,000 | Power at n = 140,000 | Power at n = 300,000 | N for 80% power | N for 90% power |
|---|---|---|---|---|---|---|---|---|
| 0.10 | 1.00 | 0.045 | 0.144 | 0.515 | 0.953 | 0.999 | 80,405 | 109,717 |
| 0.20 | 1.00 | 0.045 | 0.147 | 0.516 | 0.924 | 0.991 | 86,157 | 124,240 |
| 0.30 | 1.00 | 0.045 | 0.153 | 0.519 | 0.885 | 0.965 | 96,290 | 155,481 |
| 0.40 | 0.99 | 0.046 | 0.158 | 0.507 | 0.843 | 0.925 | 113,820 | 216,514 |
| 0.50 | 0.98 | 0.046 | 0.164 | 0.511 | 0.802 | 0.888 | 138,591 | - |
| 0.75 | 0.91 | 0.046 | 0.190 | 0.505 | 0.729 | 0.805 | 295,025 | - |
| 1.00 | 0.84 | 0.047 | 0.212 | 0.512 | 0.691 | 0.741 | - | - |
| 1.19 | 0.80 | 0.050 | 0.228 | 0.512 | 0.654 | 0.703 | - | - |
| 1.50 | 0.75 | 0.053 | 0.264 | 0.505 | 0.615 | 0.662 | - | - |
| 2.00 | 0.69 | 0.063 | 0.303 | 0.505 | 0.582 | 0.629 | - | - |

*"-": the target power was not reached within the simulated trial sizes.*
